# Meta-Analyses and Meta-Regression Analyses Revealed that Crimean-Congo Hemorrhagic Fever Disease Associates with Coagulopathy Independently of Thrombocytopenia

**DOI:** 10.64898/2026.01.24.26344481

**Authors:** Roaa Khafaji, Sura Safi, Reem S. Ubayis, Sally Rita Witwit, Eddean Witwit, Ahmed Jawad, Laurent O. Mosnier, Juan C. de la Torre, Haydar Witwit

## Abstract

Crimean Congo hemorrhagic fever (CCHF) disease, caused by CCHF virus (CCHFV), poses a significant fatality risk whose underlying pathological mechanisms, including the contribution of coagulation factors imbalances and platelets abnormalities, remain poorly understood. Here we present a meta-analysis and meta-regression analysis using clinical data from coagulation assays and platelet parameters as predictive disease indices with the goal of uncovering pathognomonic factors and to pave a path for the development of effective therapeutic approaches.

**Methods:** We systematically analyzed published studies reporting coagulation assays and platelet indices in patients with confirmed CCHF. Data from 1,779 patients across published studies were analyzed to assess associations between laboratory parameters and fatality risk, while evaluating heterogeneity and prognostic significance.

**Results:** Fatal outcomes were strongly associated with elevated liver enzymes (AST: 1116.71 ± 1454.08 IU/mL; ALT: 446.56 ± 457.41 IU/mL) and prolonged clotting times (PT: 19.53 ± 6.57 s; aPTT: 64.02 ± 23.13 s; INR: 1.53 ± 0.56). D-dimer levels did not significantly predict fatality. Thrombocytopenia and coagulopathy emerged as independent risk factors for adverse outcomes. Notably, protein C and protein S levels did not differ between survivors and non-survivors, suggesting that the coagulopathy is not purely consumptive or a result of impaired hepatic synthesis. In contrast, mildly reduced antithrombin levels (83.65 ± 19.90) were weighted toward increased mortality.

**Simple Summary:** Crimean-Congo Hemorrhagic Fever (CCHF) causes high mortality through hemorrhage, but whether this reflects thrombocytopenia alone or combined coagulopathy remains unclear. We conducted meta-analyses of studies each of coagulation parameters (PT, aPTT, INR, fibrinogen, D-dimer, protein C and protein S, antithrombin) from survivors vs non-survivors. Fatal cases showed combined thrombocytopenia and coagulopathy (elevated PT/aPTT/INR, reduced fibrinogen/antithrombin) and liver damage (elevated AST/ALT). Protein C and protein S were unaffected, suggesting complex mechanisms beyond simple consumption or hepatic failure. These findings indicate coagulopathy contributes independently of thrombocytopenia to CCHF hemorrhage, supporting combined platelet and coagulation factor replacement therapy.

## 1. Introduction

CCHF patients exhibit clinical signs of both bleeding and coagulation defects, suggesting an overlap between platelets disorder and coagulopathies such as purpura, epistaxis, heavy menstrual cycle, hemarthrosis, melena, vaginal bleeding, gingival bleeding, hematemesis, hematochezia among others. CCHF progresses in three phases: pre-hemorrhagic (fever, myalgia, nausea; 1-7 days), hemorrhagic (petechiae, epistaxis, GI bleeding; 20-30% fatality), and convalescent (recovery or multiorgan failure; overall CFR 10-70%). Standard treatment includes ribavirin (controversial efficacy) and supportive care (fluids, transfusions) [1,2]. These clinical hemorrhages suggest coagulopathy beyond thrombocytopenia.

CCHFV (genus *Orthonairovirus*, family *Nairoviridae*), the causative agent of CCHF, is an enveloped virus with a tri-segmented (L, M, and S segments) negative strand RNA genome [3–7] and causes severe and fatal illness in humans, with fatality rates that can reach more than 70% [8]. The L segment encodes the virus RNA-directed RNA polymerase (RdRp, L protein), which is responsible for directing transcription and replication of the viral genome. The L protein contains an ovarian tumor-like protease (OTU) domain that has deISGylating and deubiquitylating functions, which have been shown to counteract the host innate immune response [9,10]. The M segment encodes the virus glycoprotein precursor (GPC) that is processed by host proteases to produce a GP160/85 domain that is further processed to a mucin-like domain (MLD), GP38, the Gn and Gc glycoproteins and the medium non-structural protein (NSm) [11]. The S gene encodes the nucleoprotein (NP) in one reading frame and the small non-structural protein (NSs) in an opposite-sense open reading frame [12]. CCHF is classified into six genotypes. Type I, II and III are dominant in Africa, type IV is found in Asia, while type V and VI associated with Europe [13]. The inclusion of CCHF in the WHO R&D Blueprint priority list underscores the urgent need for the development of therapeutics against CCHF [14]. CCHFV enters host cells via clathrin-mediated endocytosis, a process that involves binding of CCHFV glycoprotein Gc to the low-density lipoprotein receptor (LDLR), which serves as a main CCHFV entry receptor in mammalian cells [15,16], while DC-SIGN facilitates this entry process [17], and with both cholesterol and endosomal low pH playing essential roles [18,19]. Also, CCHFV has a broad cell tropism [20,21].

Humans can be infected by CCHFV via direct tick bites, exposure to animal products (e.g., in slaughterhouses), and sexual contact [22,23], but nosocomial infections have also been reported [24]. The *Ixodidae* family and specifically *Hyalomminae* subfamily is the most reported tick to transmit CCHFV [25], but other genera of ticks can also carry the virus [26]. As temperatures rise and rainfall patterns shift, the habitat of ticks is expanding [27–33], allowing the hyalomma vector to live and reproduce in geographic regions that were previously outside the range of environments tolerated by the tick [33–36]. This expansion has led to an increase in the number of CCHFV cases in regions where the disease was previously rare or absent and an expansion of CCHF disease endemic regions.

Hemorrhage is a main factor contributing to the CCHF mortality rate [37,38], but the underlying mechanisms remain controversial [39–48]. Reduced platelet counts in CCHF have been reported, but it remains controversial whether bleeding associated with CCHF solely reflects thrombocytopenia [8,39,46–53]. This study addresses specific coagulation knowledge gaps through meta-analysis. Here, we present the results of a meta-analysis and meta-regression analyses of hematology laboratory parameters related to coagulation and bleeding in CCHF patients. These parameters included platelet counts, the prothrombin time (PT) and international normalized ratio (INR), as a surrogate for the extrinsic factors II, V, VII, X, the activated partial thromboplastin time (aPTT), as a surrogate for intrinsic factors XII, XI, IX, VIII, protein C and protein S as a surrogate for natural anticoagulant pathways, D-dimer as a surrogate for the fibrinolytic pathway, and antithrombin and fibrinogen as a surrogate of thrombin formation. Since the liver is the main site for the synthesis of many coagulation factors, we also investigated liver cellular pathology using alanine transferase (ALT) and aspartate transferase (AST) levels as a surrogate for liver damage. We used survivability versus fatality as a parameter to track these laboratory findings. Here, we present the results of a meta-analysis and meta-regression analyses of hematology laboratory parameters related to coagulation and bleeding in CCHF patients. These parameters included platelet counts, prothrombin time (PT), activated partial thromboplastin time (aPTT), international normalized ratio (INR), protein C and protein S, D-dimer, antithrombin, fibrinogen, and liver enzymes (ALT/AST). Survivability versus fatality was used as the outcome parameter to track associations between these laboratory findings and clinical outcomes.

## 2. Materials and Methods

### 2.1. Literature search

**Search engine and duration:** Literature searches were conducted using the PubMed database between January 2022 and May 2023. Studies published before this period were also eligible, provided they were indexed in PubMed up to May 2023. **PICO criteria**: **Population**: CCHF patients (see inclusion criteria); **Exposure**: coagulation laboratory parameters (platelets, PT, aPTT, INR, ALT, AST, fibrinogen, D-dimer, antithrombin, protein C, and protein S); **Comparator**: survivors vs. fatal cases; **Outcome**: coagulation abnormalities predicting survivability. **Search strategy:** The following terms were used: “Crimean-Congo Hemorrhagic Fever” or “CCHF” combined with one of the following keywords: prognostic, predictors, mortality, fatal, survivors, or score. In addition, clinical laboratory parameters were included in the search using the following terms: platelets, PT (prothrombin time), aPTT (activated partial thromboplastin time), INR (international normalized ratio), ALT (alanine aminotransferase), AST (aspartate aminotransferase), fibrinogen, D-dimer, antithrombin, protein C, and protein S. **Study selection process:** In the initial screening phase, review articles, systematic reviews, non-related literature, and studies not meeting basic inclusion criteria were excluded. Subsequent eligibility assessment excluded studies that did not report any of the specified laboratory parameters, lacked standard deviations (or convertible dispersion measures), or did not distinguish outcomes by survivability. **Final inclusion:** Studies were included if they reported mean values with corresponding standard deviations (or dispersion measures convertible to standard deviations) for at least one of the specified laboratory parameters and provided data stratified by survivors versus non-survivors. No additional exclusions were applied based on age group, study design (beyond the initial screening), or other clinical characteristics.

### 2.2. Data collection and analysis

A total number of studies of k=18 for platelets [8,39,46–48,52,54–65], 12 for PT [39,47,48,52,55–58,60–63], 12 for aPTT [39,47,48,52,54–57,61–63,65], 9 for INR [39,47,52,55,57,59–61,65], 5 for D-dimer [39,46,48,52,57], 7 for fibrinogen [39,46,48,52,56,62], 12 for each of AST [46–48,52,54–56,59,61–63,65] and ALT parameters [46–48,52,54–56,59,61–63,65], and 2 for antithrombin, protein C and protein S [39,52] were included in the analysis. Data were transferred into an excel file and R studio was used to conduct analyses using the following packages: meta, tidyverse, plyr, Matrix, devtools, readxl, meta, ggplot2, ggrepel, plotly, viridis, dplyr. Some of these packages were used to understand the visualization but were not used directly in this paper. Tables and plots were saved as images for final usage. The analysis was carried out using the standardized mean difference as the outcome measure. A random-effects model was applied to the data. The heterogeneity amount (i.e., 𝝉^𝟐^, stands for Tau^2^, is the estimated between-study variance in a random-effects meta-analysis, it represents the variance of the true effect sizes across studies, *θ_i_=θ+u_i_, u_i_∼N(0,τ2)* where *θ* = average true effect, uᵢ = random deviation for study I^2^, τ² = variance of those deviations), was estimated using the DerSimonian-Laird estimator [66]. In addition to the estimate of 𝝉^𝟐^, the 𝑸-test for heterogeneity [67] and the 𝑰^𝟐^ ((inconsistency, a measure of heterogeneity that is dependent on 𝑸 test and df, where I^2^ = 100% x (𝑸-df)/𝑸), where 𝑸 is Cochran’s χ² statistic and df = k – 1 (k = number of studies)) statistic [68] are reported. If heterogeneity is detected (i.e., 𝝉%^𝟐^ > 𝟎, independent of the 𝑸-test result), the true outcomes’ prediction interval is also provided [69]. Studentized residuals and Cook’s distances are utilized to investigate whether studies can be identified to be outliers and/or influential in the context of the model [70]. Studies with a studentized residual with values larger than the 𝟏𝟎𝟎 × *𝟏 − 𝟎. 𝟎𝟓/(𝟐 × 𝒌)3th percentile of a standard normal distribution are categorized as potential outliers (i.e., utilizing a Bonferroni correction with two-sided 𝜶 = 𝟎. 𝟎𝟓 for 𝒌 studies included in the meta-analysis). Studies with a Cook’s distance exceeds the median plus six times the interquartile range of the Cook’s distances are considered to be influential. The rank correlation test [71] and the regression test [72], using the standard error of the observed outcomes as predictor, are used to check for funnel plot asymmetry. The analysis was carried out using R (version 4.3.2) [73] and the metafor package (version 4.4.0) [74].

Radial (Galbraith) plots were constructed to visualize individual study influence and identify outliers contributing to heterogeneity. Each study is plotted with x-coordinate representing the standardized mean difference divided by its standard error, and y-coordinate representing the inverse of the standard error (1/SE). Studies clustering near the central regression line (within ±2 standard deviations, shown as dashed lines) exert minimal influence on the pooled estimate. Points falling outside this range indicate potential outliers or influential studies, whose removal can be tested for impact on overall results [75–77].

## 3. Results

### 3.1. Laboratory factors associated with survival in CCHF patients

To observe whether there is a significant correlation between laboratory parameters and risk of mortality, we conducted a meta-analysis using standardized mean difference between two groups, CCHF survivors and non-survivors. We investigated platelets count, PT, aPTT, INR, D-dimer, fibrinogen, AST and ALT, we found that all parameters showed significant correlation, where high platelets count (z= 7.7300, 𝑝 <0.0001) and fibrinogen levels (z= 3.1374, 𝑝 = 0.0017) favored survivors group, while high values of PT (z= −7.5581, 𝑝 <0.0001), aPTT (z= −6.0964, 𝑝 <0.0001), INR (z= −3.7529, 𝑝 =0.0002), AST (z= −5.2432, 𝑝 <0.0001) and ALT (z= −4.4142, 𝑝 <0.0001) weighted toward the non-survivors group, with the exception of D-dimer which did not show significant difference (z= −1.4245, 𝑝 =0.1543). However, with the exception of fibrinogen (Tau^2^ = 0, I^2^ = 0), the metanalyses exhibited heterogeneity. Nevertheless, the outcomes of our studies are aligning in the same direction as the estimated average outcome (Fig. 1). To examine outlier influences, a studentized residuals revealed that none of the studies had a value larger than their t-distribution, so no indication of outliers in this model with the following parameters: PT (±2.8653), aPTT (±2.8653), INR (±2.7729), D-dimer (±2.5758), and fibrinogen (±2.6383). However, one study (Gul et al 2015) [54] had a value > ±2.9913 for platelet count, one study (Parlak et al 2015) [55] had a value > ±2.8376 for AST and one study (Ali et al 2007) [56] had a value > ±2.8376 for ALT. According to the Cook’s distances, an influential analysis method developed to detect studies that significantly affect the analysis parameter upon their removal from the analysis group [78], none of the studies could be considered overly influential with exception of the following studies (with their respective laboratory parameter): Demirci et al, 2023 (fibrinogen) [46] and Parlak et al, 2015 (AST) [55], and Ali et al, 2007 (ALT) [56]. In addition, we performed a cumulative meta-analysis to observe the size effect chronologically, which validated the size difference between the two groups observed with the meta-analysis. Notably, antithrombin levels were available in only 2 studies [39,52] (n=2, I²=0%) and showed significantly reduced values in non-survivors (83.65±19.90% activity, z=-2.14, *p* =0.032). Similarly, protein C and protein S were reported in 2 studies [39,52] (n=2, I²=0% for both) and showed no significant differences between survivors and non-survivors (Figure S2). Due to the limited number of studies, these results are presented as hypothesis-generating rather than definitive findings.

**Figure 1.**
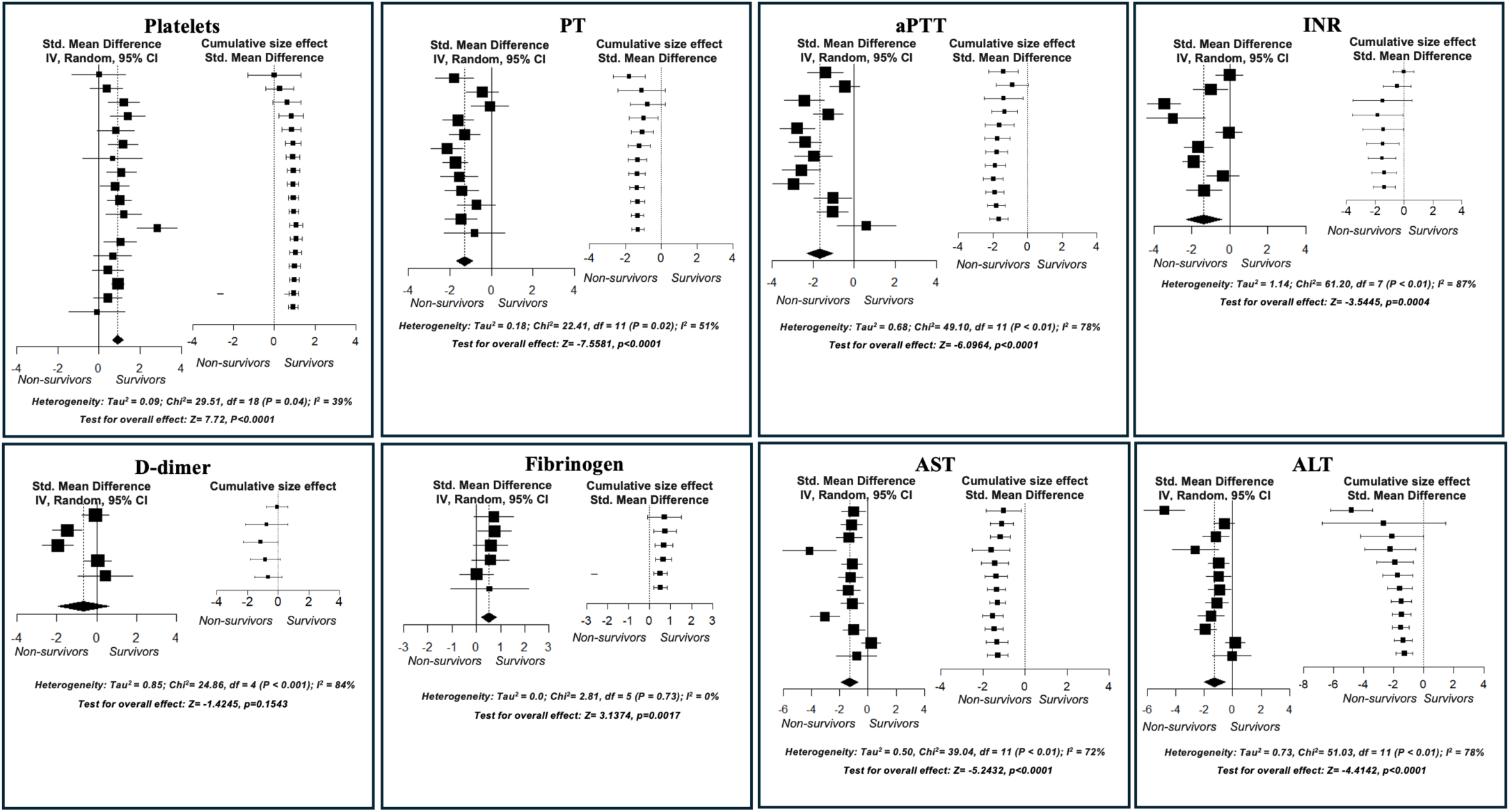
Forest plots showing the observed outcomes and random-effects model estimates. The left panel presents individual studies ordered chronologically, with the oldest study at the top and the most recent study at the bottom. Squares indicate study-specific estimates (with size proportional to sample size), and horizontal lines denote 95% confidence intervals (CI). The diamond represents the pooled effect size across all studies. The right panel illustrates the cumulative effect estimates over time, also sorted by year of publication. CI indicates confidence interval; df, degrees of freedom; IV, interval variable; PT, prothrombin time; aPTT, activated partial thromboplastin time; INR, international normalized ratio; AST, aspartate aminotransferase; ALT, alanine aminotransferase. Data were extracted and subjected to meta-analysis using RevMan model in R (meta and metafor packages).

### 3.2. Impact of individual studies on the total size effect

To understand how the significance of the individual studies affected the total size effect and the distribution around the mean and to examine symmetry bias, we used a contour funnel plot. Neither the rank correlation, a test for publication bias [71], nor the regression test [79] indicated any funnel plot asymmetry for all meta-analyses, for platelet count (𝑝 = 0.6540 and 𝑝 = 0.5512, respectively), PT (𝑝 = 0.2496 and 𝑝 = 0.2115), aPTT (𝑝 = 0.4590 and 𝑝 = 0.3907), INR (𝑝 =0.3585 and 𝑝 = 0. 1927), D-dimer (𝑝 = 0.2333 and 𝑝 = 0.4410), fibrinogen (𝑝 =1.0000 and 𝑝 =0.8227), and ALT (𝑝 =0.0311 and 𝑝 =0.0092), except for AST (𝑝 = 0.0030) where the regression test indicated a funnel plot asymmetry (𝑝 =0.0256), figure 2. This validates that the size effect is not related to bias in most of the parameters.

**Figure 2.**
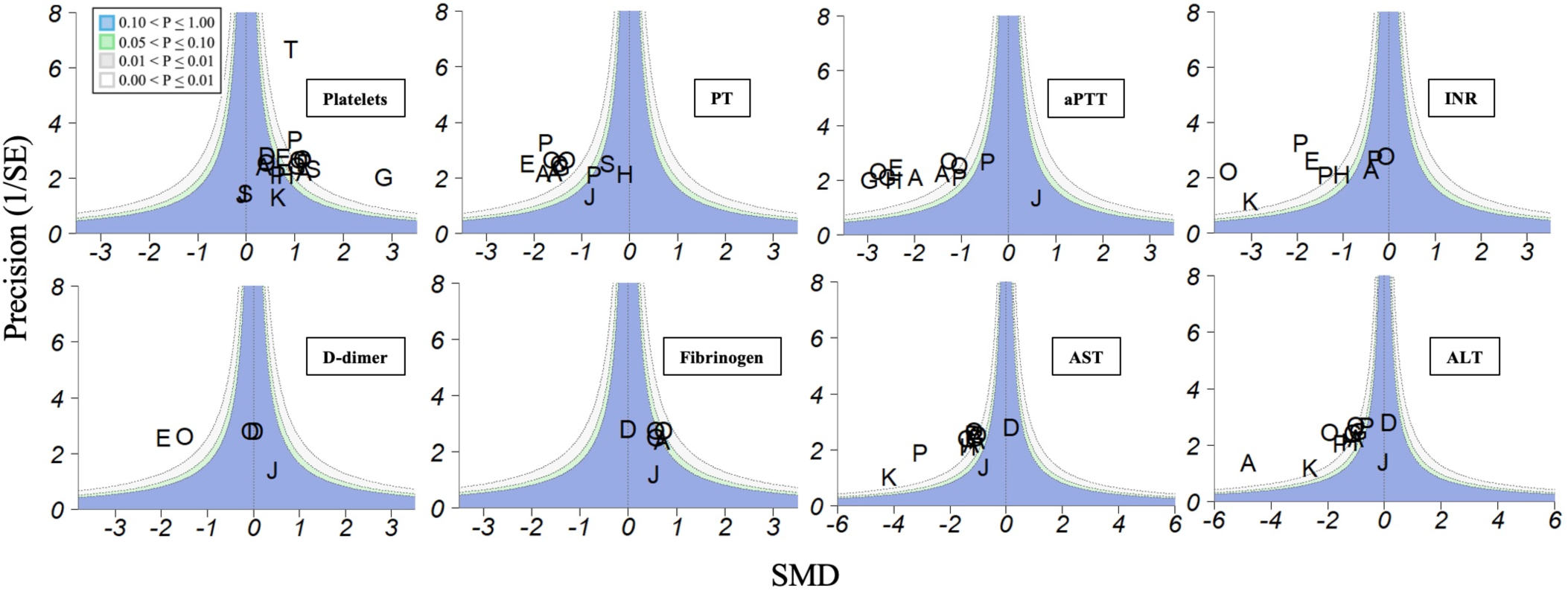
Contour-enhanced funnel plot illustrating the relationship between sampling variance and effect size, with shading indicating levels of statistical significance. The plot was used to assess potential asymmetry suggestive of publication bias. Individual studies are labeled on the plot using the first letter of the first author’s surname (letters repeated where initials overlap). Shaded regions indicate statistical significance contours (*p* < 0.1, 0.05, 0.01). A rank correlation test was performed to evaluate the association between effect size and its variance. SMD, Standardized Mean Difference; PT, prothrombin time; aPTT, activated partial thromboplastin time; INR, international normalized ratio; AST, aspartate aminotransferase; ALT, alanine aminotransferase. Data were analyzed using R (meta and metafor packages).

### 3.3. Subgrouping and meta-regression analyses based on sample size

To investigate the heterogeneity of the meta-analyses, we used sample size as a factor to subgroup studies, with 50 as the cutoff number. This resolved the heterogeneity in platelets count, where studies with sample size above 50 were homogenous (Tau^2^ and I^2^ are 0) with test of total effect of 12.21 (𝑝 < 0.01), while the heterogeneity still existed in studies with sample size less than 50, see figure 4. This approach did not resolve the heterogeneity for the rest of parameters. aPTT carried the least heterogeneity (n>50) 66%, the heterogeneity of aPTT didn’t affect the total size effect, on the other hand, one study in PT meta-regression showed no significance (Hatipoglu et al) [47], however another study, by Ekiz et al [57], while affirming the total effect, it reinforced heterogeneity due to its large size effect. For INR, the total size effect is significant, but the heterogeneity indicated by I^2^ value of 87%. AST heterogeneity caused mostly by one study (Demirci et al) [46] pulling the effect toward no significance, while another study, Parlak et al [55], affirming the significant of the size effect but creating a heterogenous effect. ALT heterogeneity stems from one study (Demirci et al) [46], weighted the effect toward no significance. Subgrouping did not affect fibrinogen, nor the D-dimer, the former remains minimally heterogenous while the latter retains heterogeneity, figure 3.

**Figure 3.**
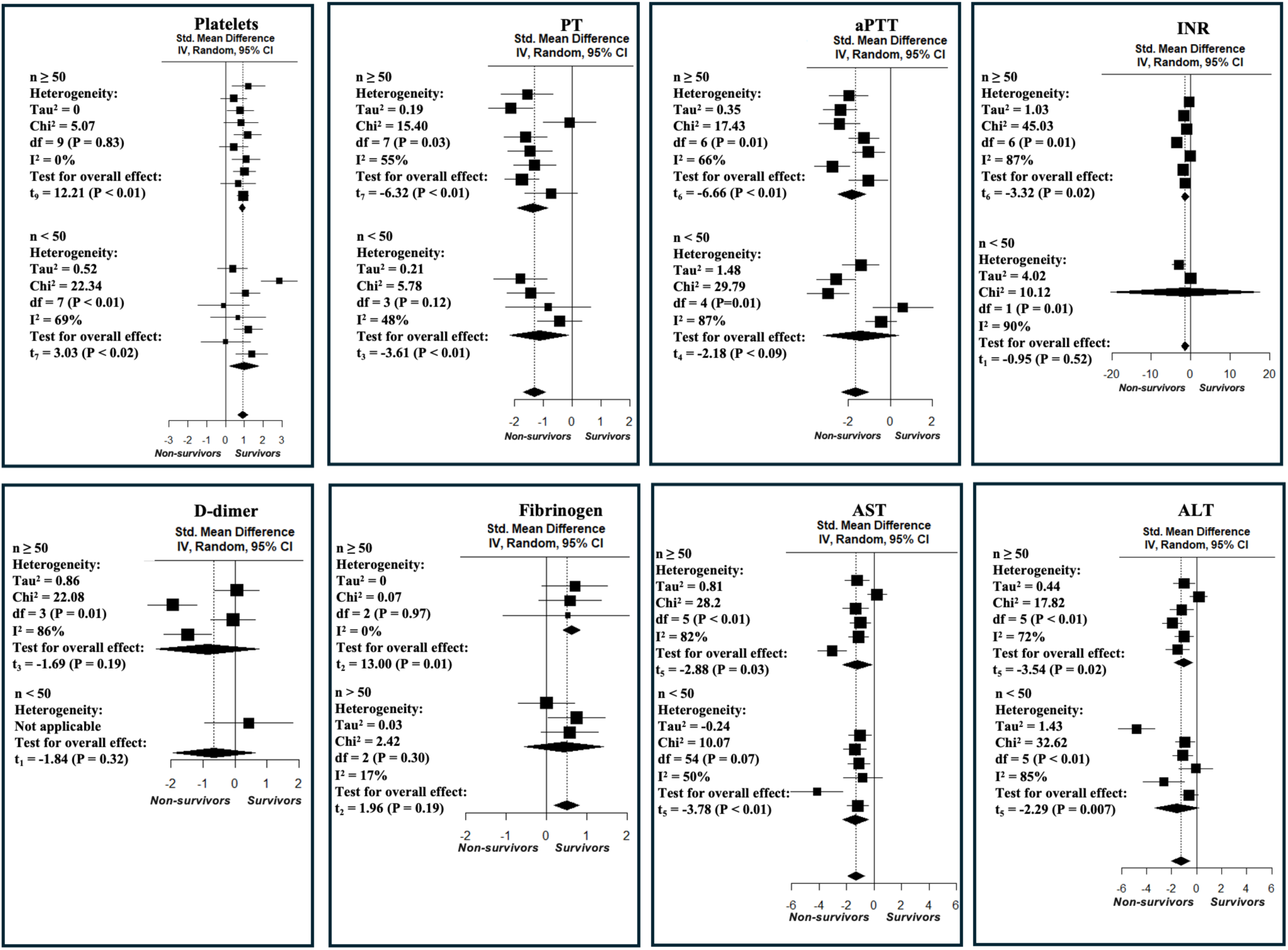
Forest plot showing the observed outcomes and random-effects model estimates based on subgroup analyses using a sample size cutoff of 50. Squares represent study-specific estimates (with size proportional to sample size), horizontal lines denote 95% confidence intervals (CI), and diamonds indicate pooled subgroup effects. CI indicates confidence interval; df, degrees of freedom; T, test for overall effect; IV, interval variable; PT, prothrombin time; aPTT, activated partial thromboplastin time; INR, international normalized ratio; AST, aspartate aminotransferase; ALT, alanine aminotransferase. Data were analyzed using R (meta and metafor packages).

**Figure 4.**
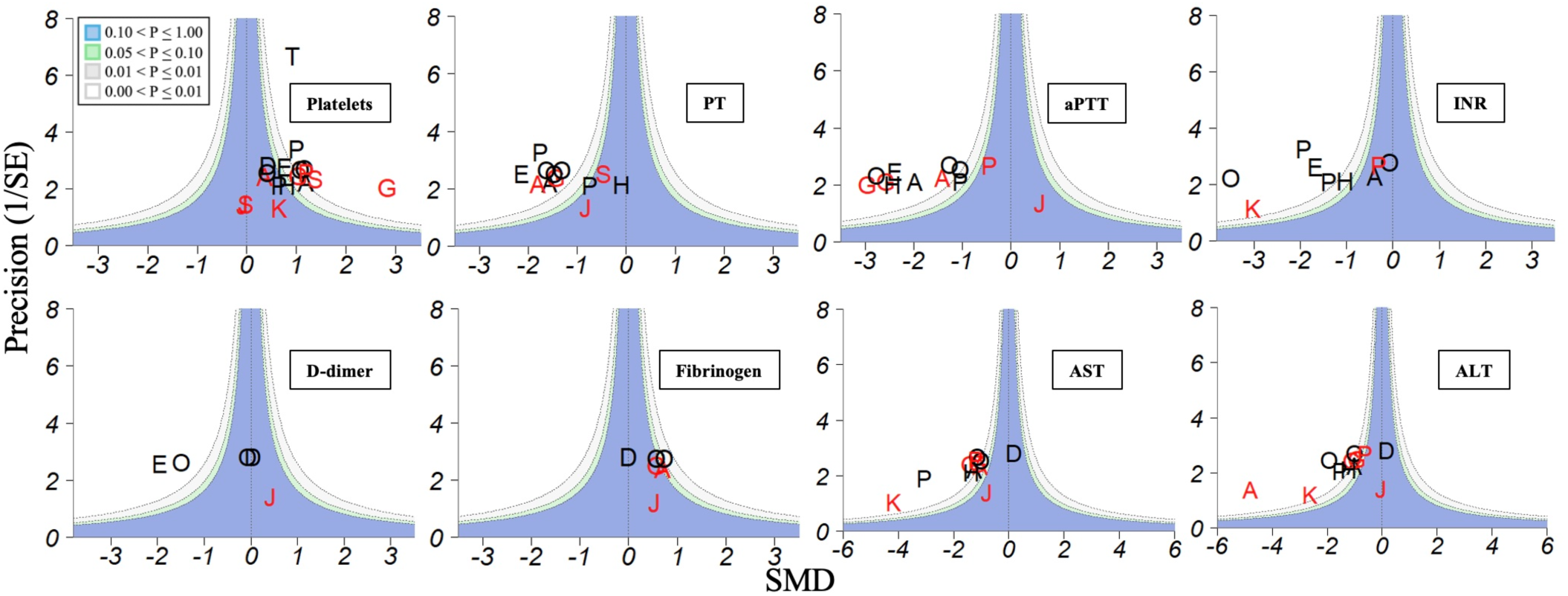
Contour-enhanced funnel plot used to assess asymmetry in sampling variance relative to effect size, with contours indicating levels of statistical significance. Individual studies are labeled on the plot using the first letter of the first author’s surname (letters repeated where initials overlap). Black color letters are studies where the n, sample size, (control group) ;: 50, red color for n < 50. Shaded regions indicate statistical significance contours (*p* < 0.1, 0.05, 0.01). A rank correlation test was performed to evaluate the association between effect size and variance. SMD, Standardized Mean Difference; PT, prothrombin time; aPTT, activated partial thromboplastin time; INR, international normalized ratio; AST, aspartate aminotransferase; ALT, alanine aminotransferase. Data were analyzed using R (meta and metafor packages).

### 3.4. Impact of individual studies on the meta-regression analysis

After filtering out studies with sample size <50, we investigated both rank correlation and regression test of the meta-regression. Studies were depicted on a funnel plot (figure 4) and neither the rank correlation nor the regression test indicated any funnel plot asymmetry, around the SMD for all parameters, for platelet count (1.0000 and 𝑝 = 0.6588, respectively), PT (𝑝 = 0.1789 and 𝑝 =0.0549), aPTT (𝑝 = 0.7726 and 𝑝 = 0.4581), INR (𝑝 = 1.0000 and 𝑝 = 0.8573), fibrinogen (𝑝 = 1.0000 and 𝑝 = 0.1783), AST (𝑝 = 0.0167 and 𝑝 = 0.0006) and ALT (*p* =0.2722 and 𝑝 = 0.1924), except for D-dimer where the regression test indicated a funnel plot asymmetry (𝑝 <0.0001) but not the rank correlation test (𝑝 = 0.0833). This affirms that the size effect is not related to bias due to funnel plot asymmetry except for D-dimer.

### 3.5. Assessment of heterogeneity and total size effect using modified Radial (Galbraith) Plot

To validate our findings and examine both size effect and sample size effect of each study on the total effect in this meta-analyses and meta-regression analyses, we devised a modified radial (Galbraith) plot to depict meta-analyses and meta-regression analyses to observe how single studies changed the gravity of the total size effect (figure 5A). We observed a similar effect of Gul et al 2015 [54] as an outlier, due to its large size effect, in platelets meta-regression, however; its effect is rather confirmatory. Both AST and ALT meta-regression analyses revealed outliers, Parlak et al 2015 [55] and Ali et al 2007 [56] respectively. Reversing the depiction, where the size of the circle is a representation of sample size and the color is a depiction of size effect (figure 5B), permitted to detect studies influencing the total effect that were visibly undetectable in figure 5A (e.g., two studies negatively contributed to the total effect but did not affect heterogeneity in the platelets meta-regression analysis). No other parameters showed outliers beyond ±2SD regression bounds.

**Figure 5.**
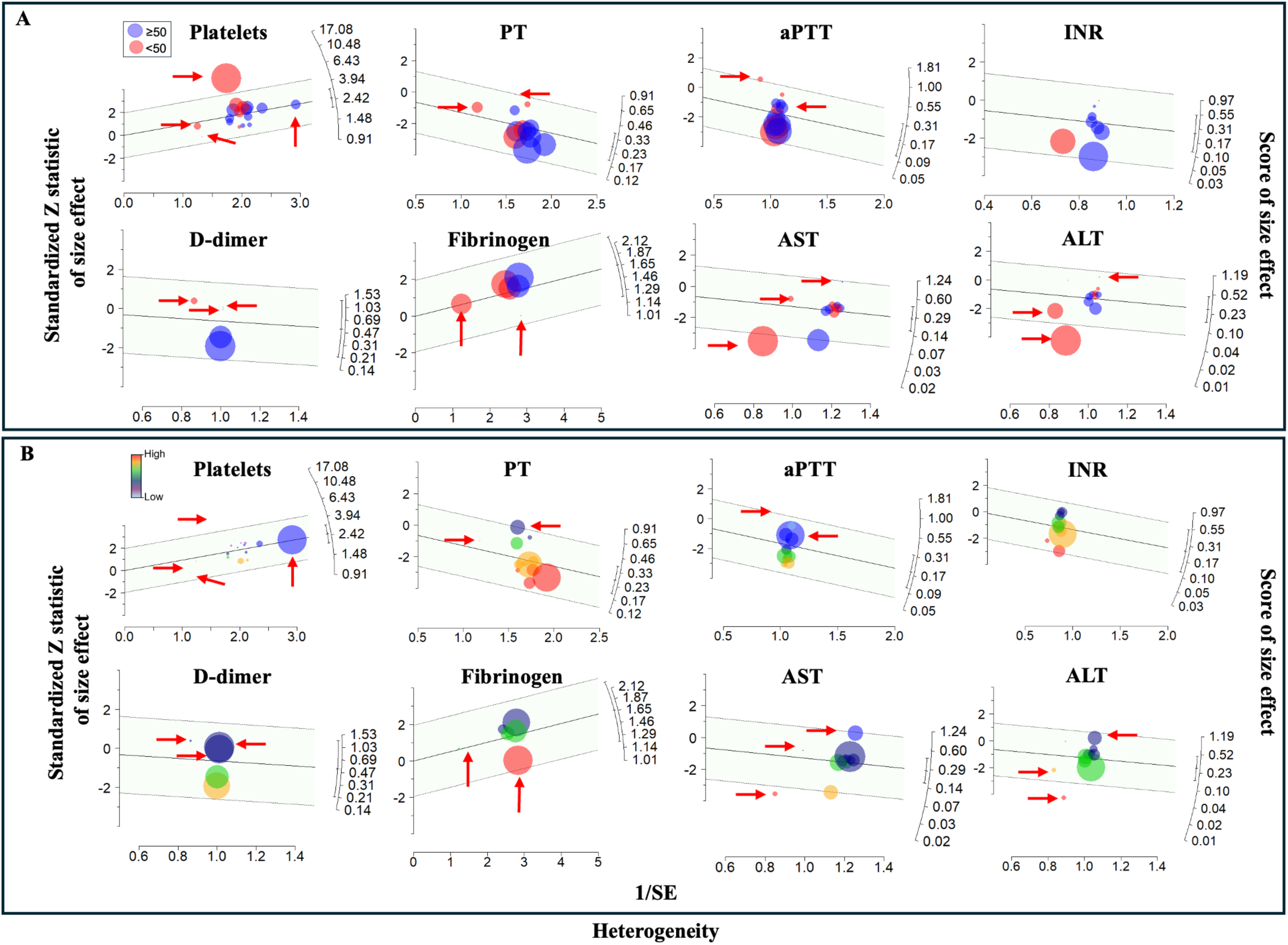
Radial (Galbraith) graphs of effect sizes for laboratory parameters. (A) radial (Galbraith) plot displays individual study-level effect estimates paramaters’ differences, modeled using a random-effects DerSimonian–Laird (DL) approach. Standardized mean differences (yi) were calculated using Hedges’ method and plotted on a radial scale with exponentiation applied for interpretability. Point size are scaled proportional to the absolute value of each study’s effect estimate using the formula:Point Size = (|yi| × 10) / max(|yi|), larger absolute effect sizes appear as larger circles. Point color reflects total sample size (c) to visualize potential small-study effects: studies with c < 50 are plotted in semi-transparent red, and studies with c ≥ 50 are plotted in semi-transparent blue. This plot allows assessment of the distribution of effect sizes, heterogeneity, and potential asymmetry in accordance with PRISMA and Cochrane reporting guidelines. (B) This radial (Galbraith) plot displays the same study SMD estimates with a focus on study influence and magnitude. The model and radial transformation are identical to Figure A. Point size are scaled proportional to the study contribution metric (c) using the formula: Point Size = (|c| × 10) / max(|c|). Studies with greater influence appear as larger circles. Point color encodes effect size magnitude using a continuous gradient ranging from red → orange → green → blue → purple. Darker hues indicate larger deviations from the null. This visualization emphasizes influential studies and provides a graphical assessment of heterogeneity, effect distribution, and directional clustering, consistent with PRISMA- and Cochrane-recommended meta-analysis visualizations. X-axis is 1/SE, y-axis is Zi= 𝜃”i/SEi. PT, prothrombin time; aPTT, activated partial thromboplastin time; INR, international normalized ratio; AST, aspartate aminotransferase; ALT, alanine aminotransferase. Data were analyzed using R (meta and metafor packages).

### 3.6. Normalized size effect of individual studies

To observe whether individual normalized cutoff values can be used as prognostic factors, normalized values of the parameters with their standard deviations were plotted for each study. We observed that all parameters lack clear cutoff values to be useful as prognostic factors due to overlapping standard deviations of the survivors and non-survivors (figure 6). This indicates that these parameters may not be utilized, alone, as predictors of CCHF fatality.

**Figure 6.**
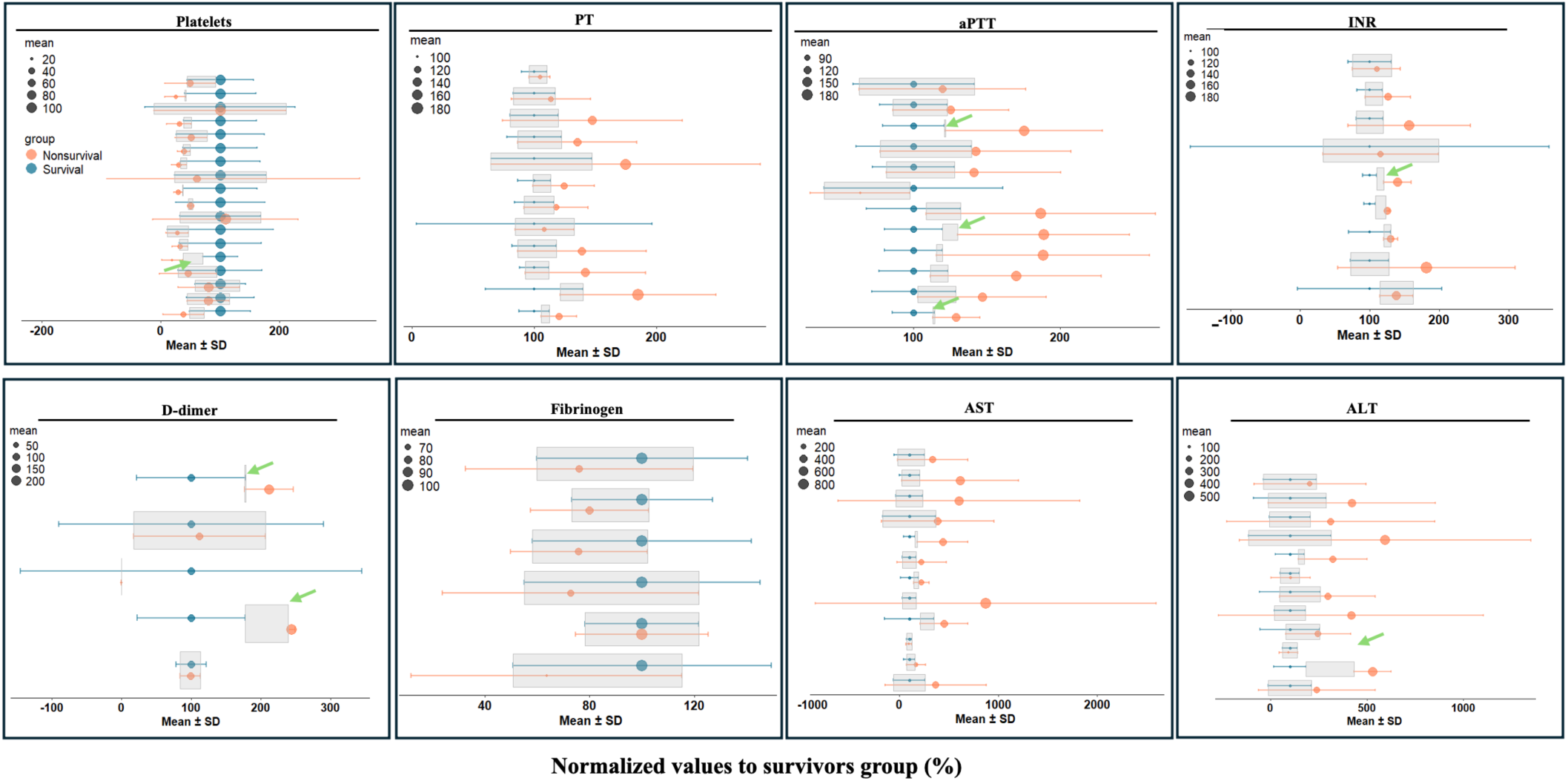
Mean differences between survivors and non-survivors. Values are normalized to the mean of the survivor group. Error bars indicate variability, and green arrows highlight studies in which error bars do not overlap. Mean ± standard deviation (SD) values for survival and non-survival groups across included studies. Horizontal error bars represent SD, and gray shaded rectangles indicate the regions of overlap between groups. The majority of studies exhibit overlapping SDs, with only a few showing distinct separation, illustrating high inter-study variability and highlighting the difficulty in defining robust cutoff values for these outcomes. PT, prothrombin time; aPTT, activated partial thromboplastin time; INR, international normalized ratio; AST, aspartate aminotransferase; ALT, alanine aminotransferase. Data were analyzed using R (meta and metafor packages).

## 4. Discussion

The etiology of hemorrhage associated with CCHFV infection is a controversial issue. Here, we utilized meta-analyses and meta-regression analyses to investigate the role of the coagulation pathway in CCHF and found that elevated values of PT, aPTT, and INR support that both the extrinsic and intrinsic coagulation pathways play a role in hemorrhage associated with fatal cases of CCHF. This is supported by some reports documenting prolongation of either PT or aPTT but not both in fatal cases of CCHF, whereas fibrinogen was affected in all fatal cases. The nature of the coagulopathy remains to be determined. Overall, our meta-analyses and meta-regression analyses are consistent with the criteria of diffuse intravascular coagulation (DIC). Elevated AST and ALT indicate that liver damage might contribute to dysregulation of coagulation factor synthesis levels in these patients. However, protein C, protein S and antithrombin (n=2 studies) [39,52] showed differential patterns: protein C and protein S unaffected while antithrombin mildly reduced (83.65±19.90% activity). These findings preclude definitive conclusions about hepatic synthetic capacity versus consumption, as coagulation factors have distinct half-lives (protein C: ∼6-8h; antithrombin: ∼2-3 days) and may be differentially affected by acute liver injury [80]. However, the much shorter half-live of protein C versus antithrombin may suggest that impaired liver synthesis is an unlikely explanation for the decrease in antithrombin but not protein C. Notably, reduced antithrombin levels in non-survivors were consistently observed across the two available studies (I²=0%). While this finding suggests potential consumptive coagulopathy or DIC-like mechanisms in severe CCHF, the limited number of contributing studies (n=2) necessitates cautious interpretation. These results should be regarded as hypothesis-generating, meriting validation in larger prospective cohorts to confirm their prognostic utility and underlying pathophysiological relevance. Available information was insufficient to delve deeper in the nature of the coagulopathy as a more detailed analysis of individual coagulation factor levels and fibrinolytic factors is lacking to date. Also, our results did not exclude that direct cell host-viral interactions could contribute to alterations in coagulation. Two key questions remain to be addressed (*1*) whether specific consumption of coagulation factors ensues during infection among fatal cases of CCHF, and/or (*2*) whether the coagulopathy is associated with the release of viral proteins and progeny and/or is ensuing the induction of cellular and/or endothelial damage.

Our study has inherent limitations due to extensive clinical and methodological variability across source studies, including: (1) timing of sample collection relative to disease onset (admission vs. peak illness), (2) sample handling protocols affecting coagulation parameters, (3) laboratory equipment specifications (different coagulometers, reagent sensitivities), (4) concomitant therapies (ribavirin, FFP), (5) patient comorbidities, and (6) viral strain differences. These sources of variation were explicitly addressed through: 1) Random-effects models using DerSimonian-Laird estimation, which provide conservative pooled estimates accounting for between-study heterogeneity (τ² reported for all analyses). 2) Comprehensive heterogeneity assessment: Q-test, I² (0-87% across parameters), and prediction intervals for all significant findings. 3) Galbraith (radial) plots identifying influential outliers (Gul 2015 platelets, Parlak 2015 AST, Ali 2007 ALT), with sensitivity analyses confirming result stability upon exclusion. 4) Subgroup analyses by sample size (n<50 vs. n≥50), which resolved platelets heterogeneity completely (τ²=0, I²=0% in larger studies). 5) No small-study effects (contour-enhanced funnel plots symmetrical except AST/D-dimer, confirmed by rank correlation and Egger’s regression). Effect directions remained remarkably consistent across studies (favoring survivors or non-survivors, never opposing), supporting the robustness of pooled estimates despite extensive clinical heterogeneity. Human genetic variation might play a role and add another layer in enhancing overlapping of margin of errors, for instance MCP-1 was shown to worsen during the course the disease [81].

Our analysis support that, in addition to thrombocytopenia, coagulopathy and liver damage play a role in CCHF-associated hemorrhage. However, we cannot rule out that the interaction of viral proteins with coagulation factors, intra or extracellularly, also contribute to CCHF hemorrhage. Current evidence does not allow us to identify which altered factor is the main driver of hemorrhage and whether the coagulopathy observed in CCHF cases represents a host adaptative response to infection, or it is specific to CCHFV pathology. For supportive therapy, platelets, fresh frozen plasma and fibrinogen should be considered to rectify the irregularities in hemostasis in CCHF. In cases of necrosis without major high-risk bleeding, low-dose heparin may be considered. It has been shown that corticosteroids are preventive in CCHF [82], and corticosteroids enhance coagulation factor and fibrinogen production [83–85]. Collectively, these pieces of evidence validate our meta-analyses that both platelets and coagulation factors play role in hemorrhage induced by CCHF disease and exposes the urgent need for more specific research in this area.

## 5. Conclusions

CCHF-related mortality is linked to combined thrombocytopenia and a distinct coagulopathy. Correction of thrombocytopenia alone may be insufficient to prevent hemorrhage; targeted therapeutic approaches addressing both platelet deficits and dysregulated coagulation are likely required to improve patient outcomes.

## Supplementary Materials

Please find the supplementary Figures at Page 16.

## Author Contributions

Conceptualization, R.K., JCT, and H.W.; Formal analysis, R.K., R.S.U., and H.W.; Investigation, R.K., R.S.U., and H.W.; Methodology, R.K. and H.W.; Project administration, R.K. and H.W.; Resources, N.A. ; Software, H.W.; Validation, R.K., R.S.U., and H.W.; Visualization, H.W.; Writing—original draft, R.K. and H.W.; Writing—review and editing, R.K., R.S.U., S.S., A.J., E.W., SR.W, L.O.M, JCT, and H.W.; Supervision, H.W. All authors have read and agreed to the published version of the manuscript.

## Institutional Review Board Statement

NA

## Informed Consent Statement

NA

## Data Availability Statement

Data included in the article.

## Data Availability

All data produced in the present work are contained in the manuscript

---

**Figure S1.**
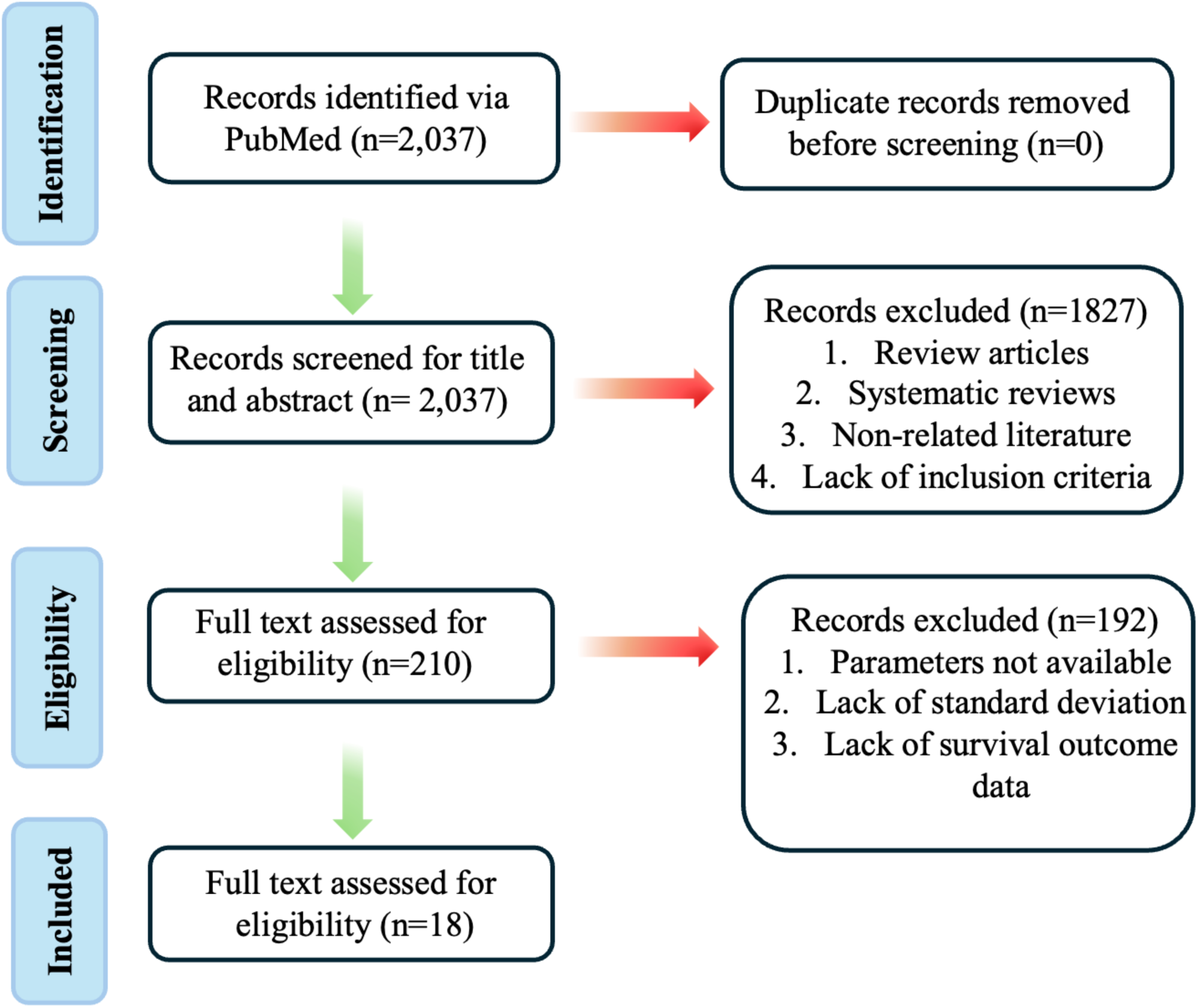
Flow chart of study selection for the systematic review and meta-analysis, showing the number of records identified, screened, excluded, and included at each stage.

**Figure S2.**
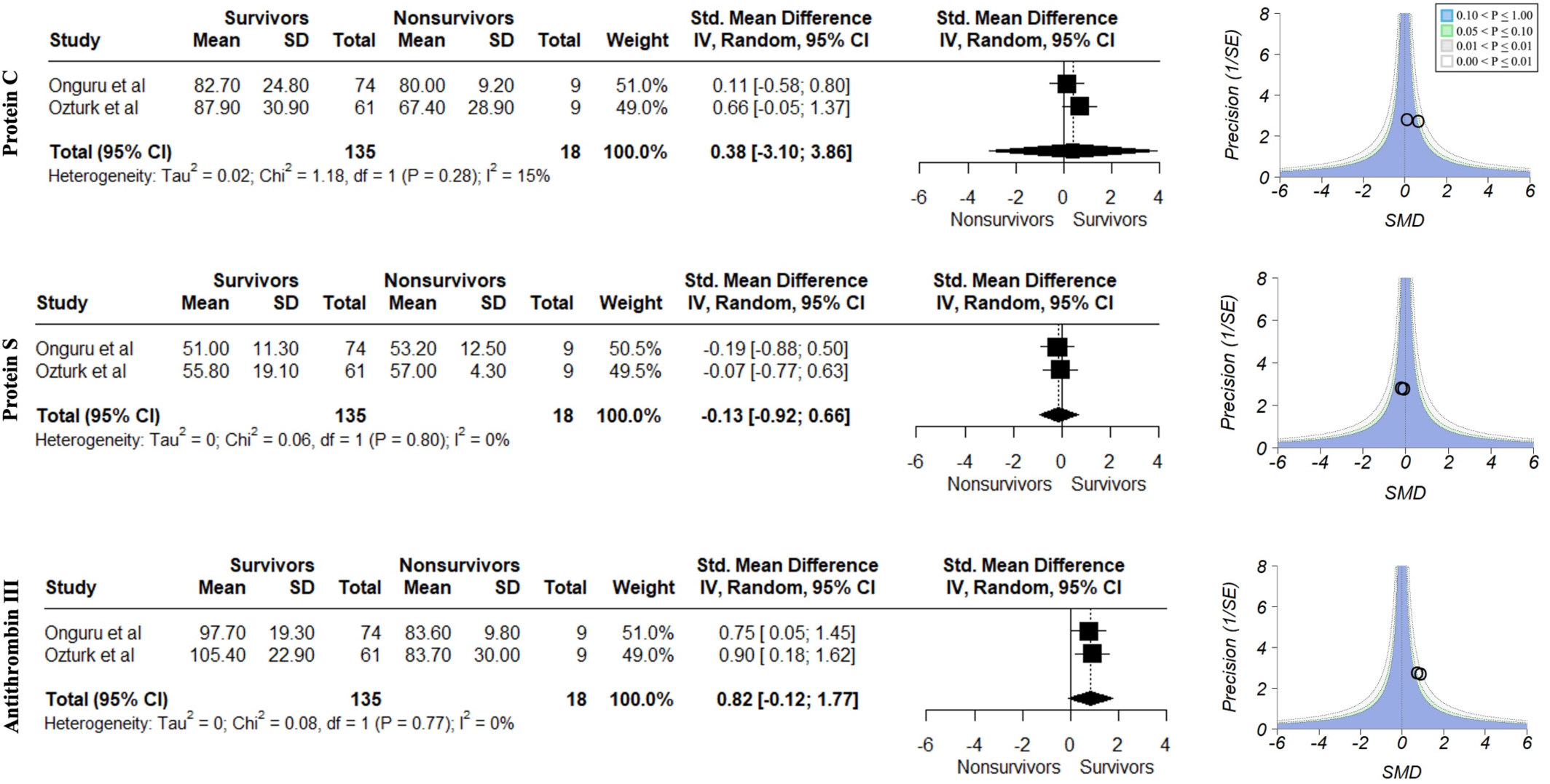
Effect of CCHF on anticoagulant protein C, protein S, and antithrombin. The figure includes forest plots showing study-specific and pooled effect estimates, alongside contour-enhanced funnel plots for each protein to assess potential asymmetry and publication bias. Data were analyzed using R (meta and metafor packages).

**Figure S3.**
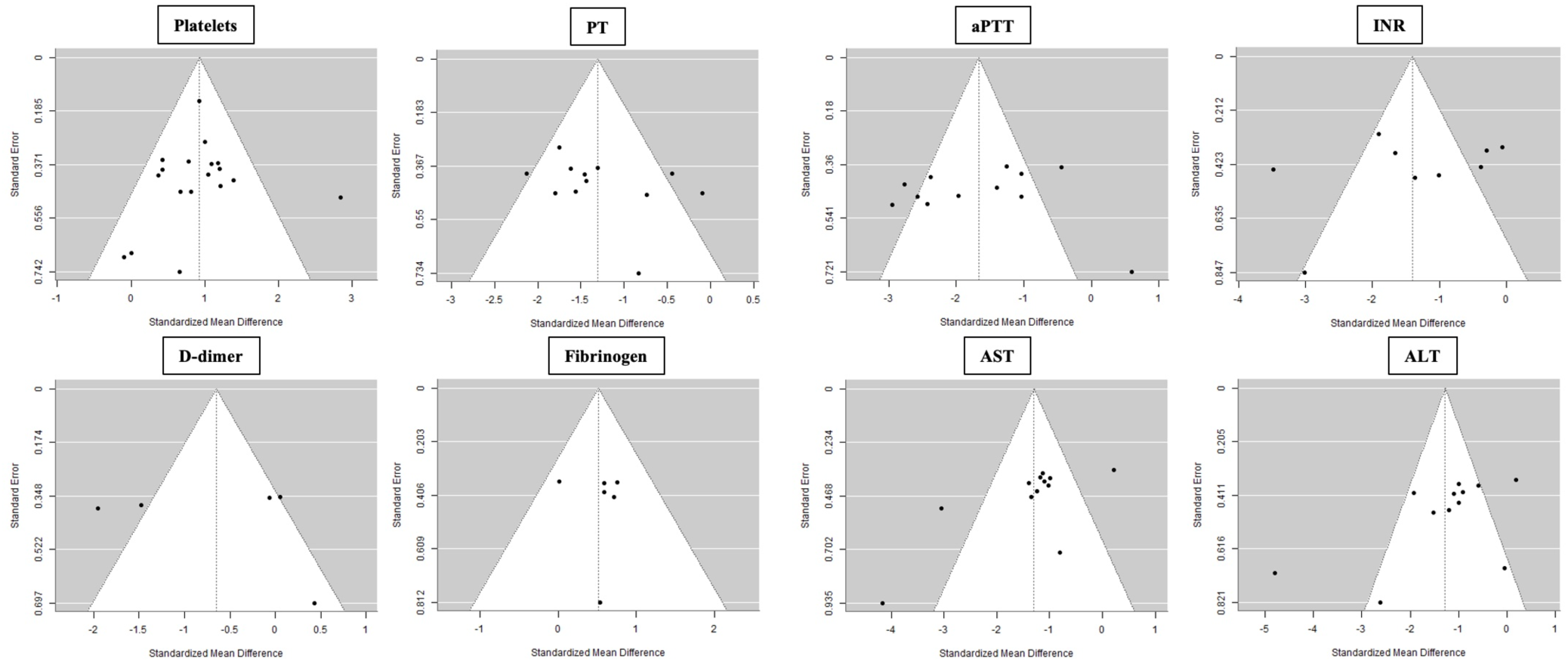
Funnel plot showing the distribution of studies to assess potential asymmetry indicative of publication bias. PT, prothrombin time; aPTT, activated partial thromboplastin time; INR, international normalized ratio; AST, aspartate aminotransferase; ALT, alanine aminotransferase. Data were analyzed using R (meta and metafor packages).

**Figure S4.**
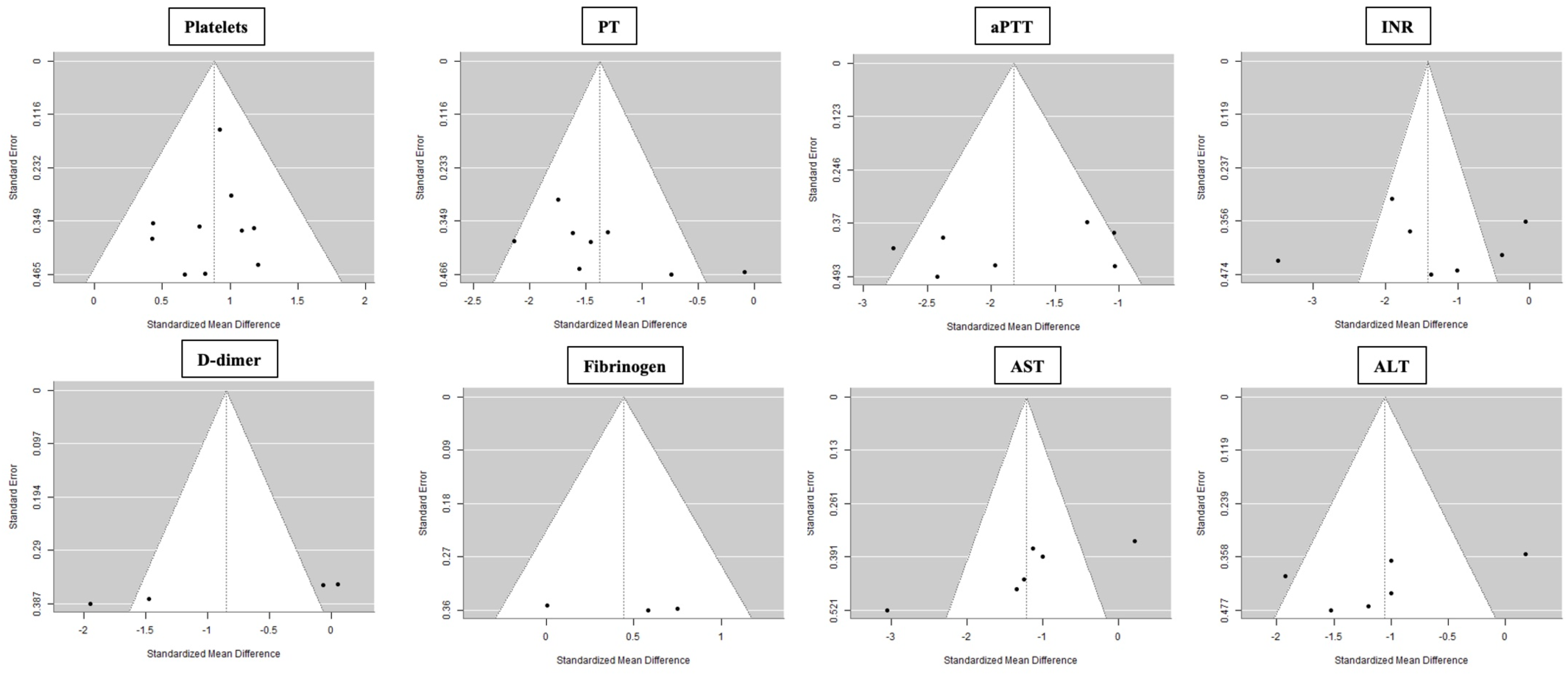
Funnel plot showing the distribution of studies with sample size >50 to assess potential asymmetry indicative of publication bias. PT, prothrombin time; aPTT, activated partial thromboplastin time; INR, international normalized ratio; AST, aspartate aminotransferase; ALT, alanine aminotransferase. Data were analyzed using R (meta and metafor packages).

